# Counties with lower insurance coverage are associated with both slower vaccine rollout and higher COVID-19 incidence across the United States

**DOI:** 10.1101/2021.03.24.21254270

**Authors:** Emily Lindemer, Mayank Choudhary, Gregory Donadio, Colin Pawlowski, Venky Soundararajan

**Affiliations:** nference, One Main Street, East Arcade, Cambridge, MA 02142, USA

## Abstract

Efficient and equitable vaccination distribution is a priority for effectively outcompeting the transmission of COVID-19 globally. A recent study from the Centers for Disease Control and Prevention (CDC) identified that US counties with high social vulnerability according to metrics such as poverty, unemployment, low income, and no high school diploma, have significantly lower rates of vaccination compared to the national average^1^. Here, we build upon this analysis to consider associations between county-level vaccination rates and 68 different demographic, socioeconomic, and environmental factors for 1,510 American counties with over 228 million individuals for which vaccination data was also available. Our analysis reveals that counties with high levels of uninsured individuals have significantly lower COVID-19 vaccination rates (Spearman correlation: −0.264), despite the fact that the CDC has mandated that all COVID-19 vaccines are free and cannot be denied to anyone based upon health insurance coverage or immigration status. Furthermore, we find that the counties with high levels of uninsured individuals tend to have the highest COVID-19 incidence rates in March 2021 relative to December 2020 (Spearman correlation: 0.388). Among the 68 factors analyzed, insurance coverage is the only factor which is highly correlated with both vaccination rate and change in COVID-19 incidence during the vaccination period (|Spearman correlation|> 0.25). We also find that counties with higher percentages of Black and Hispanic individuals have significantly lower vaccination rates (Spearman correlations: −0.128, −0.136) and lesser declines of COVID-incidence rates (Spearman correlations: 0.334, 0.330) during the vaccination period. Surprisingly however, after controlling for race, we find that the association between lack of insurance coverage and vaccination rate as well as COVID-19 incidence rates is largely driven by counties with a majority white population. Among the counties with high proportions of white residents (top 10% decile), the association between insurance coverage and vaccination rate is significant (Spearman correlation: −0.210, p-value: 0.002), but among counties with low proportions of white residents (bottom 10% decile) this association is not significant (Spearman correlation: 0.072, p-value: 0.088). Taken together, this study highlights the fact that intricate socioeconomic factors are correlated not just to COVID-19 vaccination rates, but also to COVID-19 incidence fluctuations, underscoring the need to improve COVID-19 vaccination campaigns in marginalized communities. The strong positive correlation between low levels of health insurance coverage and low vaccination rates is particularly concerning, and calls for improved public health messaging to emphasize the fact that health insurance is not required to be eligible for any of the FDA-authorized COVID-19 vaccines in the United States

## Introduction

The COVID-19 vaccine rollout in the United States has been among one of the fastest in the world^2^. However, this rapid vaccine rollout has not benefited all Americans equally, and the vaccination rate in some marginalized communities has lagged significantly behind the average^3^. It is well known that social determinants of health (SDoH) and aspects of an individual’s life that occur “outside of the four walls of healthcare” have tremendous impact on actual health status^4,5^. As we approach the status of ‘herd immunity’ with COVID-19 vaccinations, it is of critical importance that the US is not viewed as a monolith, and that the vaccination coverage to promote herd immunity is equally distributed across all communities. A recent study by the CDC showed that vaccine coverage is lower in counties with high social vulnerability based upon socioeconomic indicators (poverty, unemployment, low income, no high school diploma)^1^. This study did not, however, assess the interplay between these factors and new COVID-19 incidence rates. In addition, another recent analysis 580 US counties found that the change in COVID-19 incidence from December 1, 2020 to March 1, 2021 is significantly correlated with cumulative vaccination rate through March 1, 2021^6^. However, it remains unclear whether disparities in vaccine rollout and associated COVID-19 infection rate fluctuations have been driven by some specific socioeconomic and population health factors.

Using publicly-available data, we assessed the relationship between a large set of socioeconomic variables, vaccine coverage, and COVID-19 incidence rates in 1,510 counties across the United States. We demonstrate that variables related to uninsurance status and racial minority make-up are both negatively associated with vaccine coverage and new case incidence. By stratifying counties according to minority population percentage, however, we also find that the relationship between being uninsured and poor vaccine coverage is not uniform across all races, and that this relationship is much stronger in predominantly white counties. We additionally stratify counties by their vaccine coverage and determine the relative risk of different socioeconomic factors with high and low vaccination rates. Our findings show that the most significant risk factors for poor vaccination at the county-level are those related to poverty and environmental safety such as uninsurance prevalence, teen births, and violent crimes, and the most significant protective factors are related to food security, social connectivity, and education level.

Taken together, our findings suggest several calls to action that could be implemented to increase vaccine coverage across the United States. The first is compounded by the findings of others, and relates to the racial and disparities in vaccine coverage across the country. The second call to action, however, is novel and suggests that communication surrounding vaccination eligibility in the uninsured population, and particularly in the white uninsured population, may increase vaccination rates and decrease new case incidence.

## Methods

We analyzed 1,510 counties in the United States which have cumulative vaccination data available through March 1, 2021. These counties include over 228 million individuals from 21 states and the District of Columbia, including: California, Connecticut, Florida, Georgia, Illinois, Indiana, Maine, Maryland, Michigan, Minnesota, Missouri, Nevada, New Jersey, New York, Ohio, Oregon, Pennsylvania, Tennessee, Texas, Washington, Wyoming. A map of states included in the analysis is provided in **Supplementary Figure S1**. For counties in these states, we obtained vaccination data from state government websites (county-level vaccination data was not readily available for the other states). For each county, the *vaccination rate* was defined as the percentage of individuals in the county with at least one dose of an FDA-authorized COVID-19 vaccine as of March 1, 2021. In addition, county-level COVID-19 incidence data was obtained from the CDC COVID Data Tracker^7^. The *change in COVID-19 incidence* is defined as the 7-day rolling average COVID-19 incidence rate on March 1, 2021 minus the 7-day rolling average COVID-19 incidence rate on December 1, 2020, where the COVID-19 incidence rate is the number of new COVID-19 cases reported in the county per 10,000 individuals.

Demographic and socioeconomic data for each county was obtained from the *2020 County Health Rankings*^*8*^ resource provided by the County Health Rankings & Roadmaps program at the University of Wisconsin Population Health Institute. We applied a 75% completeness requirement to each of our variables, leaving us with 68 out of a total of 131 variables from the 2020 County Health Rankings. We note that most of the variables with limited data availability were race-specific variables for minority populations (e.g. Number of firearm fatalities − Black, Motor vehicle crash deaths - Hispanic), so these could not be included in our analysis. A complete list of demographic and socioeconomic variables with data available is included in **Table 1**. For each of the 68 county-level features, we compute the Spearman rank correlations between: (1) the feature of interest and county-level *vaccination rate* and (2) the feature of interest and county-level *change in COVID-19 incidence*. Spearman rank correlations and corresponding p-values were computed using the SciPy package^9^ in Python.

**Table 1.** Complete list of Spearman rank correlation for county-level features with vaccination rates and change in COVID-19 incidence rates. The county-level vaccination rate is defined as the percentage of individuals in the county with at least one COVID-19 vaccine dose as of March 1, 2021. The county-level COVID-19 incidence rate increase is defined as the 7-day rolling average COVID-19 incidence rate (Number of new COVID-19 cases / total population) in the county on March 1, 2021 minus the 7-day rolling average COVID-19 incidence rate in the county on December 1, 2020. For each county-level feature, we show the Spearman rank correlation coefficients for the feature vs. vaccination rate and the feature vs. COVID-19 incidence rate increase, along with the associated p-values. Rows are sorted by correlation with vaccination rate.

| County-level feature | Vaccination rate correlation | Vaccination rate p-value | COVID-19 incidence rate increase correlation | COVID-19 incidence rate increase p-value |
| --- | --- | --- | --- | --- |
| Uninsured adults | -0.264 | 0 | 0.388 | 0 |
| No highschool diploma | -0.258 | 0 | 0.199 | 0 |
| Uninsured | -0.252 | 0 | 0.381 | 0 |
| % below 18 years of age | -0.220 | 0 | -0.083 | 0.00889 |
| Teen births | -0.215 | 0 | 0.068 | 0.30242 |
| Unemployment 2020 | -0.208 | 0 | 0.008 | 0.2042 |
| Motor vehicle crash deaths | -0.200 | 0.00053 | 0.071 | 0.00263 |
| Children eligible for free or reduced price lunch | -0.199 | 0 | 0.275 | 0 |
| Uninsured children | -0.185 | 5.00E-05 | 0.282 | 0 |
| Firearm fatalities | -0.171 | 6.00E-05 | 0.072 | 0.03812 |
| Hispanic | -0.136 | 0.00537 | 0.330 | 3.00E-05 |
| % Hispanic 2020 | -0.136 | 0.00593 | 0.329 | 2.00E-05 |
| % not proficient in English | -0.131 | 0.00137 | 0.297 | 0.00125 |
| Black | -0.128 | 0.00785 | 0.334 | 0 |
| % Black 2020 | -0.128 | 0.00848 | 0.337 | 0 |
| % Poverty 2019 | -0.119 | 0.01003 | 0.187 | 0 |
| Low birthweight | -0.116 | 0.01228 | 0.160 | 0 |
| % Native Hawaiian / Other Pacific Islander 2020 | -0.115 | 0.00435 | 0.258 | 0.00069 |
| Highschool diploma only | -0.104 | 6.00E-05 | -0.203 | 0 |
| Motor vehicle crash deaths (White) | -0.100 | 0.07358 | -0.064 | 0.83054 |
| Violent crime | -0.091 | 0.02956 | 0.145 | 0 |
| HIV prevalence | -0.083 | 0.17095 | 0.396 | 0 |
| High school graduation 2020 | -0.082 | 0 | -0.158 | 0.00061 |
| Severe housing problems | -0.079 | 0.00407 | 0.422 | 0 |
| Suicides (White) | -0.075 | 0.86492 | -0.098 | 0.00548 |
| Male | -0.073 | 0.0083 | -0.157 | 0.05345 |
| % American Indian 2020 | -0.067 | 0 | 0.171 | 0.05516 |
| Population density | -0.059 | 0.30587 | 0.066 | 0.10268 |
| Suicides | -0.052 | 0.4909 | -0.062 | 0.02429 |
| Premature death rate | -0.052 | 0.289 | 0.033 | 0.11681 |
| Premature age adjusted mortality | -0.052 | 0.289 | 0.033 | 0.11681 |
| population | -0.042 | 0.19727 | 0.091 | 0.00128 |
| Driving alone to work | -0.039 | 0.28562 | -0.147 | 0.0218 |
| Sexually transmitted infections | -0.037 | 0.61559 | 0.187 | 0 |
| % Rural | -0.031 | 0.39361 | -0.033 | 0.20672 |
| American Indian | -0.018 | 0 | 0.030 | 0.01005 |
| Limited access to healthy foods | -0.011 | 0.00024 | 0.044 | 0.98005 |
| Income inequality | -0.003 | 0.36555 | 0.288 | 0 |
| Severe housing cost burden | -0.003 | 0.34655 | 0.351 | 0 |
| Percentage of households with high housing costs | 0.000 | 0.24206 | 0.348 | 0 |
| Unemployment rate 2019 | 0.002 | 0.75953 | -0.040 | 0.28671 |
| Injury deaths | 0.003 | 0.01048 | -0.088 | 0.00277 |
| Premature age-adjusted mortality (White) | 0.008 | 0 | -0.068 | 0.57323 |
| Premature death (White) | 0.009 | 1.00E-05 | -0.065 | 0.37807 |
| Children in single parent household | 0.012 | 0.40365 | 0.159 | 0 |
| % Asian 2020 | 0.016 | 0.98348 | 0.213 | 1.00E-05 |
| Injury deaths (White) | 0.018 | 0 | -0.083 | 0.44154 |
| Driving alone to works (White) | 0.023 | 0 | -0.067 | 0.58831 |
| Homeownership | 0.023 | 0.25837 | -0.177 | 0 |
| Alcohol-impaired driving deaths numerator | 0.023 | 0.00446 | -0.069 | 0.00473 |
| Food environment index | 0.024 | 0 | -0.045 | 0.20876 |
| Low birthweight (White) | 0.031 | 1.00E-05 | -0.073 | 0.44631 |
| Asian | 0.036 | 0.8999 | 0.193 | 2.00E-05 |
| Median household income 2019 | 0.045 | 0.77121 | -0.056 | 0.43362 |
| Land area | 0.061 | 0.0013 | 0.073 | 0.0245 |
| College or associates degree | 0.066 | 0.01624 | -0.138 | 0 |
| Female | 0.073 | 0.0083 | 0.157 | 0.05345 |
| White | 0.116 | 0.00458 | -0.420 | 0 |
| % White 2020 | 0.116 | 0.00493 | -0.421 | 0 |
| Not Hispanic | 0.136 | 0.00537 | -0.330 | 3.00E-05 |
| Social associations | 0.155 | 8.00E-05 | -0.335 | 0 |
| Access to exercise opportunities | 0.183 | 0 | -0.007 | 0.14089 |
| % 65 and older | 0.190 | 0 | -0.042 | 0.69076 |
| Bachelor or higher degree | 0.213 | 0 | 0.066 | 0.00066 |
| Dentists | 0.213 | 0 | -0.050 | 0.98508 |
| Some college 2020 | 0.237 | 0 | -0.112 | 0.00067 |
| Other primary care providers | 0.263 | 0 | -0.059 | 0.94591 |
| Primary care physicians | 0.286 | 0 | 0.004 | 0.39993 |

Next, we grouped each county into quartiles based on percent vaccinated through March 1, 2021. For a select number of county-level features, we computed the rates in the top and bottom quartiles, and reported the relative risks and Fisher exact test p-values. For the relative risk values, 95% confidence intervals were computed using a delta-method approximation^10^. The results are presented in **Table 2**. Finally, to disentangle relationships between race, insurance status, and our outcome variables, we isolated the top and bottom deciles of counties as determined by the percentage of their populations that were white. Within these two deciles, we performed further correlation analyses with insurance status and vaccine coverage.

**Table 2.** Comparison of county-level features in the top and bottom quartiles of vaccinated counties. County-level vaccination rate is defined as the percentage of individuals in the county with at least one COVID-19 vaccine dose as of March 1, 2021. Counties in the top quartile have vaccination rates greater than or equal to 9.21%, and counties in the bottom quartile have vaccination rates less than or equal to 5.52%. Rows have been sorted by relative risk in increasing order.

| County-level feature | Rate in top quartile of vaccinated counties | Rate in bottom quartile of vaccinated counties | Relative risk [95% CI] | Fisher exact test p-value |
| --- | --- | --- | --- | --- |
| Homicides | 0.004 | 0.007 | 0.608 [0.593, 0.623] | <0.001 |
| % Black | 10.322 | 16.262 | 0.635 [0.634, 0.635] | <0.001 |
| % Hispanic | 14.247 | 22.199 | 0.642 [0.641, 0.642] | <0.001 |
| Teen births | 1.78 | 2.573 | 0.692 [0.688, 0.696] | <0.001 |
| Disconnected youth | 5.703 | 7.942 | 0.718 [0.713, 0.723] | <0.001 |
| Uninsured adults | 9.929 | 13.375 | 0.742 [0.741, 0.743] | <0.001 |
| Firearm fatalities | 0.009 | 0.012 | 0.762 [0.748, 0.777] | <0.001 |
| Violent crime | 0.327 | 0.422 | 0.774 [0.769, 0.78] | <0.001 |
| Uninsured children | 4.348 | 5.508 | 0.789 [0.786, 0.792] | <0.001 |
| Children eligible for free or reduced price lunch | 46.206 | 57.523 | 0.803 [0.802, 0.804] | <0.001 |
| Sexually transmitted infections | 0.489 | 0.568 | 0.861 [0.856, 0.866] | <0.001 |
| HIV prevalence | 0.374 | 0.433 | 0.863 [0.856, 0.87] | <0.001 |
| Child mortality | 0.045 | 0.052 | 0.869 [0.851, 0.888] | <0.001 |
| Unemployment | 3.76 | 4.323 | 0.87 [0.867, 0.872] | <0.001 |
| % below 18 years of age | 20.698 | 23.803 | 0.87 [0.869, 0.87] | <0.001 |
| Infant mortality | 0.537 | 0.605 | 0.888 [0.87, 0.906] | <0.001 |
| % American Indian | 0.94 | 1.044 | 0.9 [0.896, 0.904] | <0.001 |
| Children in single-parent households | 31.7 | 34.864 | 0.909 [0.908, 0.91] | <0.001 |
| Severe housing problems | 17.547 | 19.23 | 0.912 [0.911, 0.914] | <0.001 |
| Low birthweight | 7.821 | 8.271 | 0.946 [0.941, 0.95] | <0.001 |
| Premature death | 0.401 | 0.421 | 0.953 [0.949, 0.956] | <0.001 |
| Severe housing cost burden | 15.014 | 15.641 | 0.96 [0.958, 0.961] | <0.001 |
| % Rural | 18.736 | 19.426 | 0.964 [0.964, 0.965] | <0.001 |
| Alcohol-impaired driving deaths | 26.017 | 26.464 | 0.983 [0.954, 1.014] | 0.274 |
| Injury deaths | 0.067 | 0.067 | 0.999 [0.992, 1.006] | 0.815 |
| Drug overdose deaths | 0.021 | 0.02 | 1.011 [0.993, 1.028] | 0.231 |
| High school graduation | 85.926 | 84.326 | 1.019 [1.017, 1.021] | <0.001 |
| Driving alone to work | 74.744 | 73.192 | 1.021 [1.021, 1.022] | <0.001 |
| Suicides | 0.014 | 0.013 | 1.038 [1.021, 1.055] | <0.001 |
| Homeownership | 64.628 | 62.024 | 1.042 [1.041, 1.043] | <0.001 |
| % Hawaiian/Pacific Islander | 0.194 | 0.186 | 1.045 [1.035, 1.055] | <0.001 |
| Access to exercise opportunities | 87.527 | 81.531 | 1.074 [1.073, 1.074] | <0.001 |
| Some college | 69.846 | 61.094 | 1.143 [1.143, 1.144] | <0.001 |
| Food environment index | 70.245 | 59.247 | 1.186 [0.884, 1.581] | 0.293 |
| % Asian | 5.948 | 4.963 | 1.198 [1.196, 1.201] | <0.001 |
| Social associations | 0.098 | 0.081 | 1.211 [1.194, 1.228] | <0.001 |
| % White | 66.837 | 54.423 | 1.228 [1.228, 1.229] | <0.001 |
| % 65 and older | 18.137 | 14.687 | 1.235 [1.234, 1.236] | <0.001 |
| Dentists | 0.082 | 0.057 | 1.44 [1.417, 1.463] | <0.001 |
| Other primary care providers | 0.125 | 0.081 | 1.532 [1.512, 1.553] | <0.001 |
| Primary care physicians | 0.094 | 0.060 | 1.579 [1.554, 1.603] | <0.001 |

## Results

Results from our correlation analyses are synthesized together in **Figure 1**. Each socioeconomic variable is plotted to show the strength of its relationship with county-level vaccine coverage (x-axis) and county-level new COVID-19 incidence (y-axis). The upper left quadrant contains variables that are associated with both increased incidence and poor vaccine coverage, and the bottom right quadrant contains variables that are associated with decreased incidence and better vaccine coverage. Inter-variable correlations are shown in **Supplementary Figure 2**.

**Figure 1.**
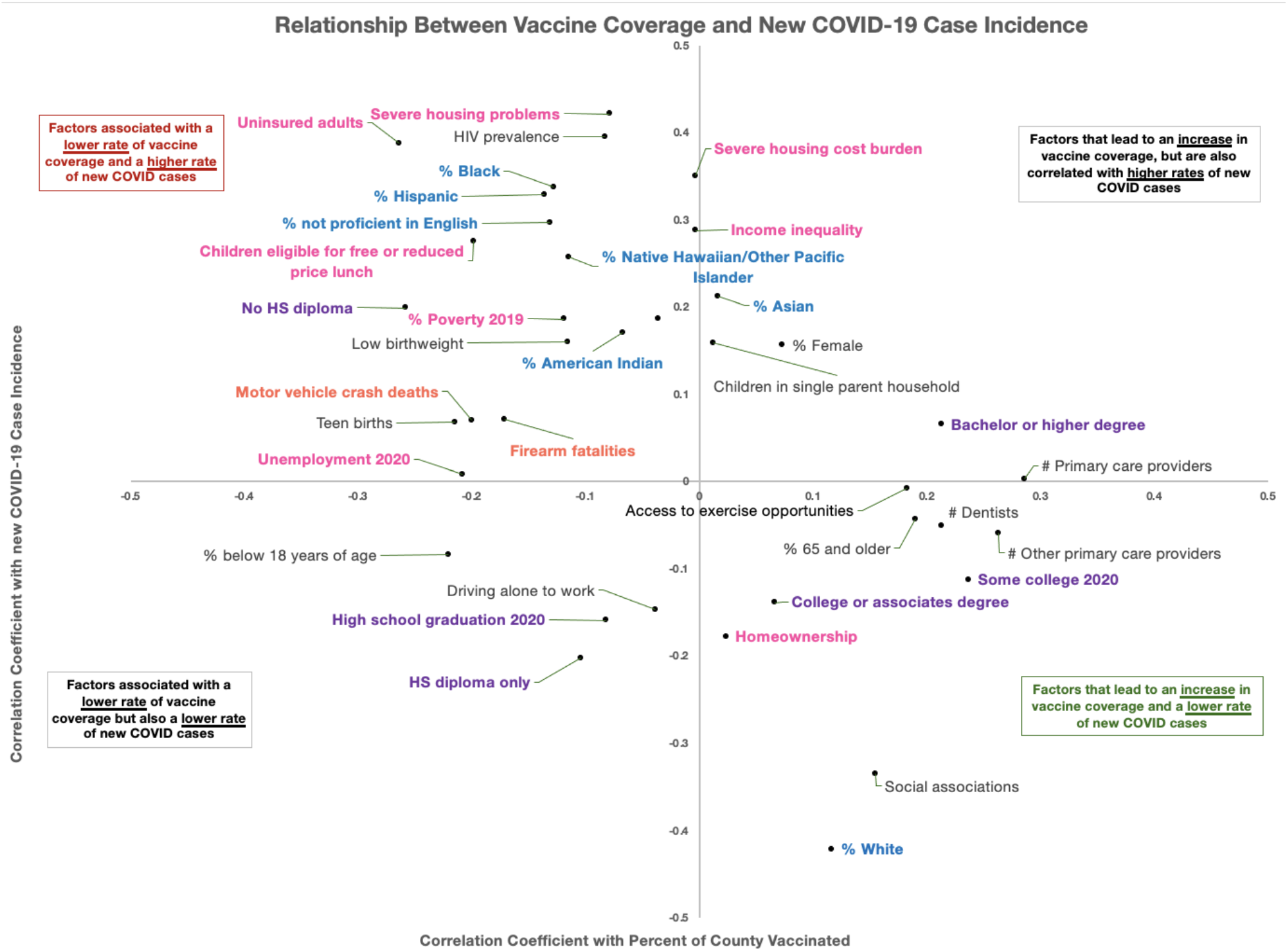
Relationship between county-level vaccine coverage and change in COVID-19 incidence rate for county-level features. On the x-axis, we show the Spearman rank correlation between the county-level feature and cumulative vaccination rate (percent of individuals in the county with 1+ vaccine dose as of March 1, 2021). On the y-axis, we show the Spearman rank correlation between the county-level feature and the change in COVID-19 incidence rate (defined as the 7-day rolling average COVID-19 incidence rate on March 1, 2021 minus the 7-day rolling average COVID-19 incidence rate on December 1, 2020. Factors are only shown here if their Spearman coefficient is greater than 0.1 along at least one dimension. Factors in pink are related to housing and income, factors in orange are related to environmental risk, factors in purple are related to education level, and factors in blue are related to race.

**Figure 2.**
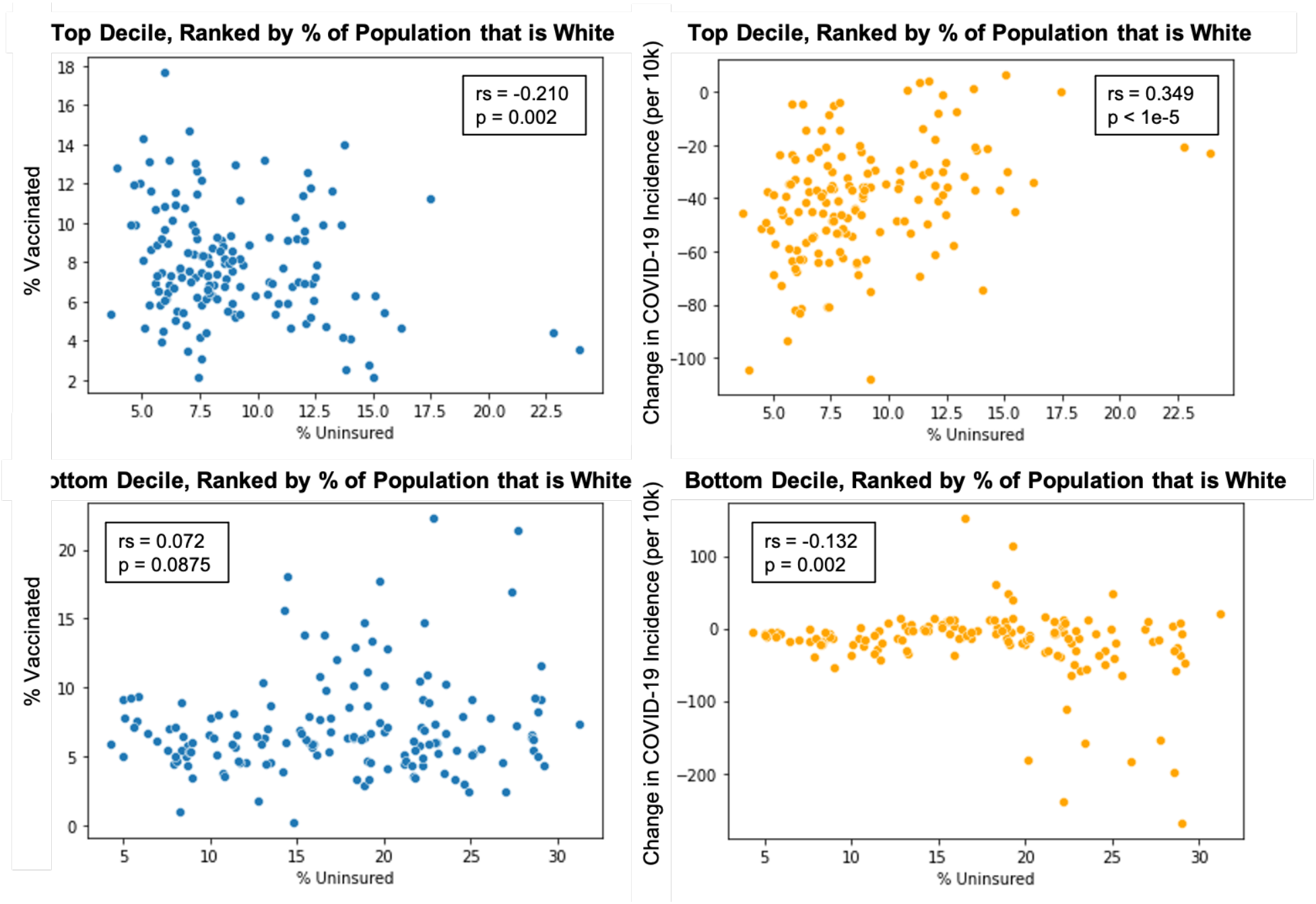
Differences in the relationship with uninsurance status between predominantly white and non-white counties. The top row shows the top decile of counties ranked by percent of the population that is white, the bottom row is the bottom decile ranked by percent white. Figures on the left show the correlation between the percent of the population that is uninsured and vaccine coverage, and figures on the right show the correlation between the percent of the population that is uninsured and new COVID-19 case incidence (r_s_ = Spearman correlation coefficient, p = significance level).

### Counties with high rates of uninsured individuals have lower vaccination rates and lower declines in COVID-19 incidence

A county’s percentage of uninsured adults and unemployed adults were both significantly correlated with the percentage of the county that had been vaccinated (Spearman correlation: −0.264, p-value: <1e-5; Spearman correlation: −0.208, p-value: <1e-5) (see **Table 1**). However, the relationship between these two factors and the incidence change in COVID-19 cases from December 1, 2020 to March 1, 2021 was vastly different. There was a significant positive correlation between the percent of the county who was uninsured and the case incidence change (Spearman correlation: 0.388, p-value: <1e-5), but no significant relationship was observed between unemployment level and new cases (Spearman correlation: 0.008, p-value: 0.20).

**Table 2** demonstrates that the relative risk related to the percent of uninsured adults in the population is 0.742, where counties in the top quartile of vaccine coverage have 9.9% uninsured adults and those in the bottom quartile have 13.4%. This translates to counties in the top quartile of vaccination coverage have a 25% lower uninsured population compared to those in the bottom quartile. **Table 3** looks at the top and bottom deciles of counties, ranked by percent of the county that is uninsured. On average, counties in the bottom decile have an uninsurance rate of 24.1%. We find that less-insured counties are more rural, have more minorities, and higher percentages of young people. The states contributing to these counties are Florida, Georgia, Indiana, Ohio, Missouri, and Texas. The bottom 25 counties for “uninsured rate” are detailed in **Supplementary Table 1**.

**Table 3.** General characteristics all counties and counties with the highest and lowest levels of insurance coverage. In the first column (Overall), we show the characteristics for all 1,510 counties with vaccination data available. In the second column (Top 10%), we show the characteristics for counties with the fewest uninsured individuals per capita (≤ 5.55%). In the third column (Bottom 10%), we show the characteristics for counties with the most uninsured individuals per capita (≥ 20.82%). Information on state, county population, major town/city, cumulative vaccination till date, and increase in COVID-incidence as of March 1, 2021 relative to December 1, 2020 are provided for each group of counties. States with at least one county in the bottom 10% based on insurance coverage are highlighted in **red**.

| Characteristic | Overall | Counties with the highest rates of insurance coverage (Top 10%) | Counties with the lowest rates of insurance coverage (Bottom 10%) |
| --- | --- | --- | --- |
| **Number of counties** | 1,510 | 151 | 151 |
| **Insurance coverage**<br>- Insured<br>- Uninsured | 88.1%<br>11.9% | 95.1%<br>4.9% | 75.9%<br>24.1% |
| **Cumulative vaccination rate (1+ dose) through March 1, 2020** | 7.7% | 8.7% | 6.8% |
| **Change in COVID-19 incidence (cases per 10K) from December 1, 2020 to March 1, 2021** | -26.8 | -34.8 | -19.0 |
| **Population**<br>- Mean<br>- Std. deviation<br>- IQR | 151,165<br>443,466<br>(15,166, 103,877) | 206,416<br>312,718<br>(36,767, 208,021) | 90,077<br>447,373<br>(4,879, 37,542) |
| **County-type**<br>- Rural<br>- Urban | 53.2%<br>46.8% | 38.7%<br>61.3% | 58.6%<br>41.4% |
| **State**<br>California<br>Connecticut<br>District of Columbia<br>Florida<br>Georgia<br>Illinois<br>Indiana<br>Maine<br>Maryland<br>Michigan<br>Minnesota<br>Missouri<br>Nevada<br>New Jersey<br>New York | California (57)<br>Connecticut (8)<br>District of Columbia (1)<br>Florida (67)<br>Georgia (159)<br>Illinois (102)<br>Indiana (92)<br>Maine (16)<br>Maryland (24)<br>Michigan (83)<br>Minnesota (87)<br>Missouri (114)<br>Nevada (15)<br>New Jersey (21)<br>New York (62) | California (7)<br>Connecticut (5)<br>District of Columbia (1)<br>Florida (0)<br>Georgia (0)<br>Illinois (24)<br>Indiana (0)<br>Maine (0)<br>Maryland (8)<br>Michigan (11)<br>Minnesota (37)<br>Missouri (0)<br>Nevada (0)<br>New Jersey (3)<br>New York (38) | California (0)<br>Connecticut (0)<br>District of Columbia (0)<br>**Florida (5)**<br>**Georgia (8)**<br>Illinois (0)<br>**Indiana (1)**<br>Maine (0)<br>Maryland (0)<br>Michigan (0)<br>Minnesota (0)<br>**Missouri (1)**<br>Nevada (0)<br>New Jersey (0)<br>New York (0) |
| Ohio<br>Oregon<br>Pennsylvania<br>Tennessee<br>Texas<br>Washington<br>Wyoming | Ohio (88)<br>Oregon (36)<br>Pennsylvania (67)<br>Tennessee (95)<br>Texas (254)<br>Washington (39)<br>Wyoming (23) | Ohio (6)<br>Oregon (0)<br>Pennsylvania (9)<br>Tennessee (0)<br>Texas (0)<br>Washington (2)<br>Wyoming (0) | Ohio (1)<br>Oregon (0)<br>Pennsylvania (0)<br>Tennessee (0)<br>Texas (135)<br>Washington (0)<br>Wyoming (0) |
| **Age**<br>- < 18 years old<br>- 18-64 years old<br>- ≥ 65 years old | 21.9%<br>59.0%<br>19.1% | 21.4%<br>60.8%<br>17.8% | 24.4%<br>57.2%<br>18.4% |
| **Gender**<br>- Male<br>- Female | 50.2%<br>49.8% | 49.9%<br>50.1% | 51.1%<br>48.9% |
| **Race**<br>- Black<br>- White<br>- Asian<br>- American Indian | 7.7%<br>75.7%<br>1.8%<br>0.7% | 5.2%<br>83.7%<br>3.2%<br>0.4% | 5.1%<br>53.3%<br>0.9%<br>0.6% |
| **Ethnicity**<br>- Hispanic<br>- Not Hispanic | 12.3%<br>87.7% | 5.6%<br>94.4% | 39.0%<br>61.0% |

### The relationship between race and insurance status is varied in its correlation with vaccine coverage and new case incidence

All racial minorities included in our analysis were correlated with higher incidence of new COVID-19 cases. With the exception of Asians, the prevalence of all other racial minority groups in counties was also negatively correlated with vaccine coverage. We compared the top 10% of counties based on the percentage of the population that was white to the bottom 10% of white counties, and performed correlation analyses in each between uninsurance status, vaccination coverage, and new case incidence. We found that in the top white counties, uninsurance status was significantly negatively correlated with vaccination coverage (Spearman correlation: −0.210, p-value: 0.002) and positively associated with new case incidence (Spearman correlation: 0.349, p-value: <1e-5). Conversely, in the bottom white counties, uninsurance rate was not significantly associated with vaccination coverage (Spearman correlation: 0.072, p-value: 0.0875) and was negatively correlated with new case incidence (Spearman correlation: −0.132, p-value: 0.002) (**Figure 2**). The bottom white decile had a racial make-up of 31% white, 17% Black, 46% Hispanic, 4% Asian, and less than 1% Native American Indian.

### Environmental risk factors and education levels are differentially correlated with vaccination rates and COVID-19 incidence

All education variables associated with higher levels of education (college or associate degree, bachelor’s degree or higher, and some college 2020) were positively correlated with vaccine coverage, and their correlations with new COVID-19 incidence rates were weak. The two variables associated with lower education levels, however (no HS diploma and HS diploma only) were both negatively correlated with vaccine coverage (**Table 1**).

Our analysis highlights two key factors (annual incidence of motor vehicle crash deaths and incidence of firearm fatalities) that we have grouped as “environmental risk factors” were negatively correlated with vaccine coverage, and positively correlated with new COVID-19. case incidence. A related correlation of note is “access to exercise opportunities” which can be viewed as an environmental protective factor, and was positively correlated with vaccine coverage, but unrelated to new COVID-19 incidence rates.

Relative risks are shown in **Table 2** for other environmental protective factors such as social associations and food environment index, which translate to higher vaccination coverage counties having a 21% and an 18% increase in each factor, respectively. In particular, counties with relatively high social association ranking, as reflected by the number of their civic organizations, have significantly higher levels of vaccination and steeper declines in COVID-19 incidence rates (**Table 1**).

## Discussion

Our findings indicate that there are several groups of socioeconomic factors that are related to the percentage of a county that is vaccinated, as well as the incidence of new COVID-19 cases, but these relationships have different strengths and directionalities, highlighting the complexities of socioeconomic disparities in the United States and the vastness of their impacts. The most striking finding from our analyses was the strong relationship with health insurance status and both vaccination coverage as well as new COVID-19 incidence rates. Going further, we found that these relationships were strongest in predominantly white counties, indicating a specific relationship between being white and uninsured and poor vaccination coverage. We hypothesize that this relationship may be due to widespread misunderstanding about individuals’ eligibility and financial responsibility for the COVID-19 vaccine, and suggest that better nationwide communication about the role of health insurance in vaccine eligibility may help vaccinate more individuals to bring us closer to herd immunity.

Despite the US government’s financial sponsorship of the COVID-19 vaccine^11^, we see a strong relationship between county-level health insurance status, percent of the county that has been vaccinated, and the incidence of new cases since the beginning of 2021. In fact, the percent of individuals uninsured in a county was the only variable whose Spearman coefficient exceeded 0.25 for both vaccine coverage and new case incidence. Juxtaposed with the relationships seen with unemployment, which was only significantly correlated with vaccination coverage but not with new COVID-19 incidence, this is an important insight. The relationship between unemployment and vaccine coverage was recently analyzed by the CDC, but not its association with new incidence rates, which we find here to be insignificant. One hypothesis to explain these differing relationships with uninsured status and unemployment is in community education about COVID-19 and vaccine eligibility. Many individuals receive information about general health and also about their vaccine eligibility status from their primary care provider^12^. This relationship does not exist in the case of individuals without health insurance, and so individuals without health insurance may be significantly uninformed about basic COVID-19 precautions that could prevent new incidences, and also about their eligibility for the vaccine. Direct messaging from the government to inform individuals that they are eligible for the vaccine regardless of insurance status may have a significant impact on both vaccine coverage and new case incidence rates.

In this study we also highlight the significant racial disparities that exist in the US in the context of COVID-19, which have been covered by numerous other studies^13–15^. Many of these studies have shown that overall outcomes of COVID-19 are poorer in communities with large populations of racial minorities. In the same vein, we demonstrate that counties with higher percentages of racial minorities are showing higher incidence rates in the first several months of 2021. While still significant, the relationship between racial minority make-up and lower vaccine coverage is weaker. These findings suggest that communities with larger populations of racial minorities are still suffering from the pandemic two-fold: the vaccine is not as readily available to them, and to those who the vaccine has not yet been administered, social precautions to prevent the spread of COVID-19 are not being as widely implemented. Interestingly, the impact of uninsurance status in minority communities is not related to vaccine coverage, and this relationship appears to be most present in predominantly white communities. We do see relationships between uninsurance and new case incidence in both predominantly white and non-white counties, although the relationships are opposing in directionality. It is difficult to draw conclusions about the cause of new cases, as the virus’ timeline varies widely across different regions of the country, but we hypothesize that it could be due to uninsured white individuals living in more rural areas with less virus spread, versus uninsured minorities living in more densely populated areas **(Supplementary Figure S4)**.

As stated in our Methods section, only 68 of the original 131 variables were able to be utilized due to limitations with data availability. We note that many variables that were lacking in complete data were those at the intersection of racial minority status and other socioeconomic factors, such as homicide rates within specific racial segments. Specifically, 52 of the 63 incomplete variables were specific to racial minority groups, and all data variables with less than 35% completeness were specific to racial minority groups. Had this been available, we may have been able to parse out more specific relationships of COVID-19, vaccination coverage, and racial minorities. We also note that one of the challenges in assessing both vaccine coverage as well as new incidence rates is in the diversity of state roll-out plans, in terms of timeline and eligibility criteria. A future retrospective analysis comparing individual states is an important next step to be taken when more data has been collected across the nation.

It is well-established that factors pertaining to race, wealth, housing and education status are tightly intertwined when it comes to healthcare^16,17^. To this end, it is not surprising that we see similar trends with lower education, poorer housing status, income inequality, and racial minorities that move in the same direction in our analyses. All of these factors show some relationship with poorer vaccine coverage, and higher recent incidence rates. We highlight in our results that there are also environmental risk factors that fall into the same pattern. In general, our findings show that all factors that fall into the bottom-right quadrant of **Figure 1**, the quadrant with the most favorable outcomes, pertain to having a higher education, a higher-paying career, homeownership, general quality of life, social connectivity and being white. For the most part, factors that fall into the top right and bottom left quadrants with mixed outcomes, are educational factors signifying a mid-range level of education, or pertain to age-related factors that directly impact vaccine eligibility such as being under 18 years old. Taken together, our findings reiterate what is known about the vast racial and socioeconomic disparities that are still ongoing in the US surrounding the COVID-19 pandemic. We find, however, an important and yet-unmentioned relationship between insurance status and vaccine coverage that we feel is addressable through better communication.

## Data Availability

After publication, the data will be made available to others upon reasonable requests to the corresponding authors. A proposal with a detailed description of study objectives and the statistical analysis plan will be needed for evaluation of the reasonability of requests.

## Supplementary Material

**Supplementary Figure S1.**
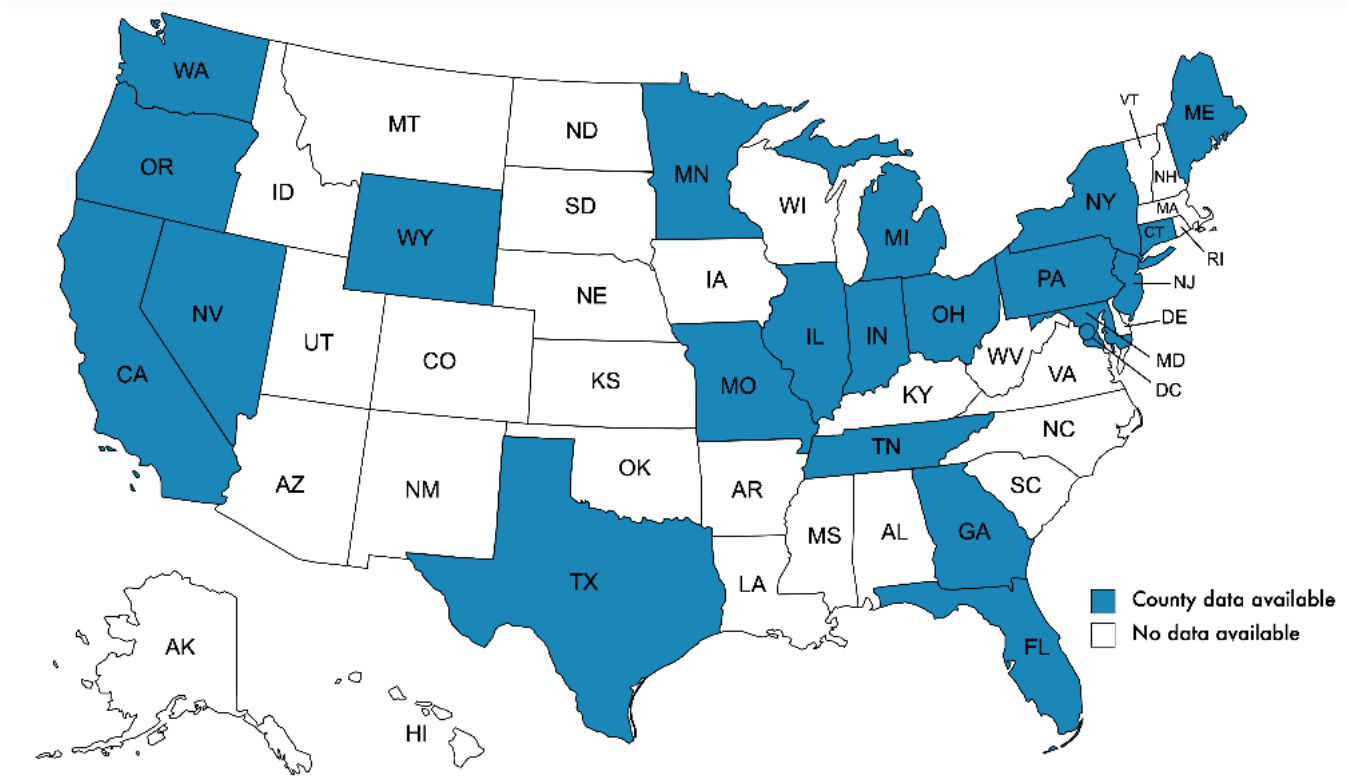
Map of the states in the USA with county-level vaccination data available.

**Supplementary Figure S2.**
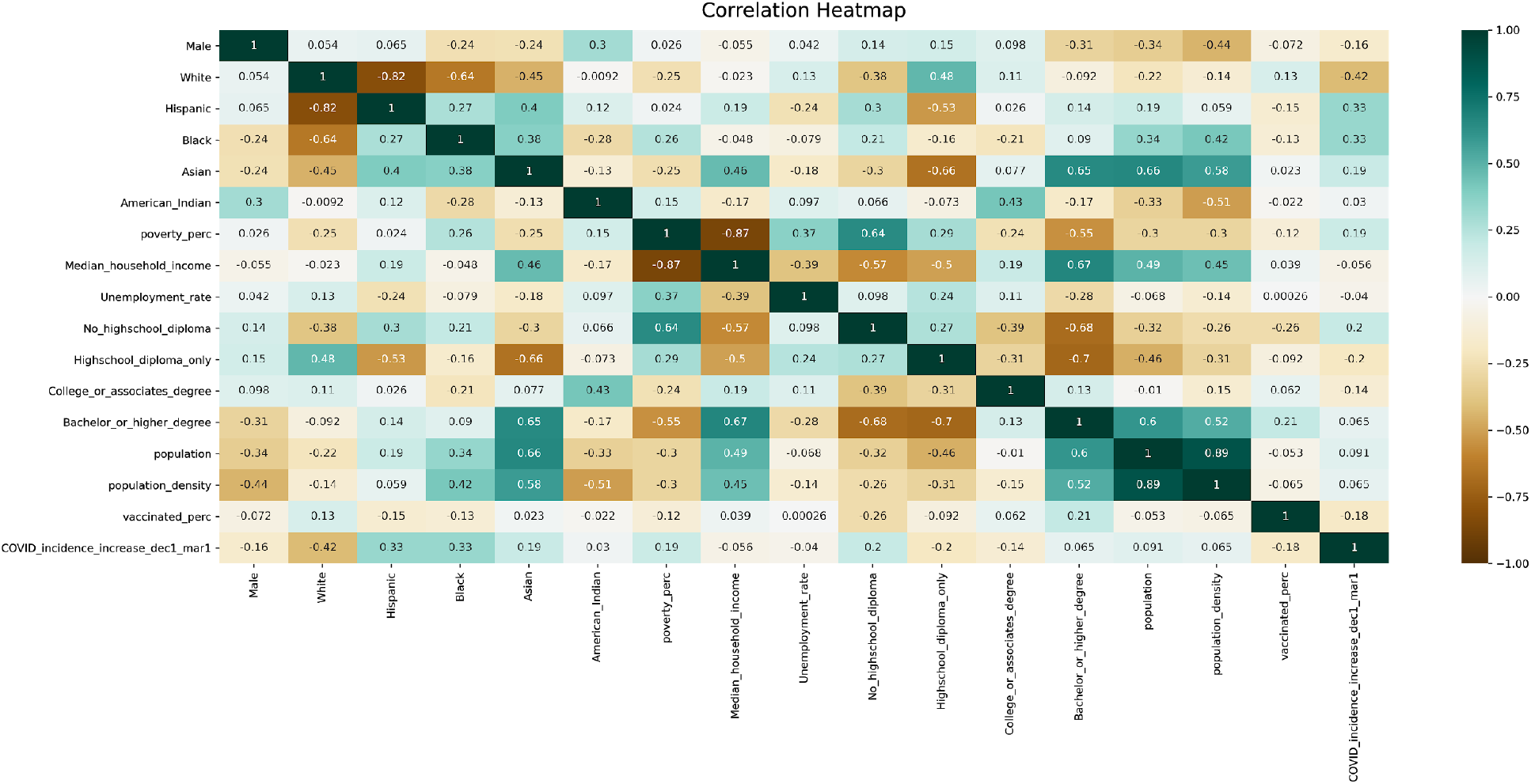
Heatmap of Spearman’s rank correlation for selected county-level features. For each pair of features, Spearman rank correlation = +1 (**dark green**) corresponds to a perfectly monotonically increasing relationship, and Spearman rank correlation = −1 (**dark orange**) corresponds to a perfectly monotonically decreasing relationship. Correlation values are calculated based upon data available from the 1,510 counties in the study dataset, and features for each county include: demographic and socioeconomic variables, cumulative vaccinated percentage through March 1, 2021, and COVID-19 increase in incidence from December 1, 2020 to March 1, 2021.

**Supplementary Figure S3.**
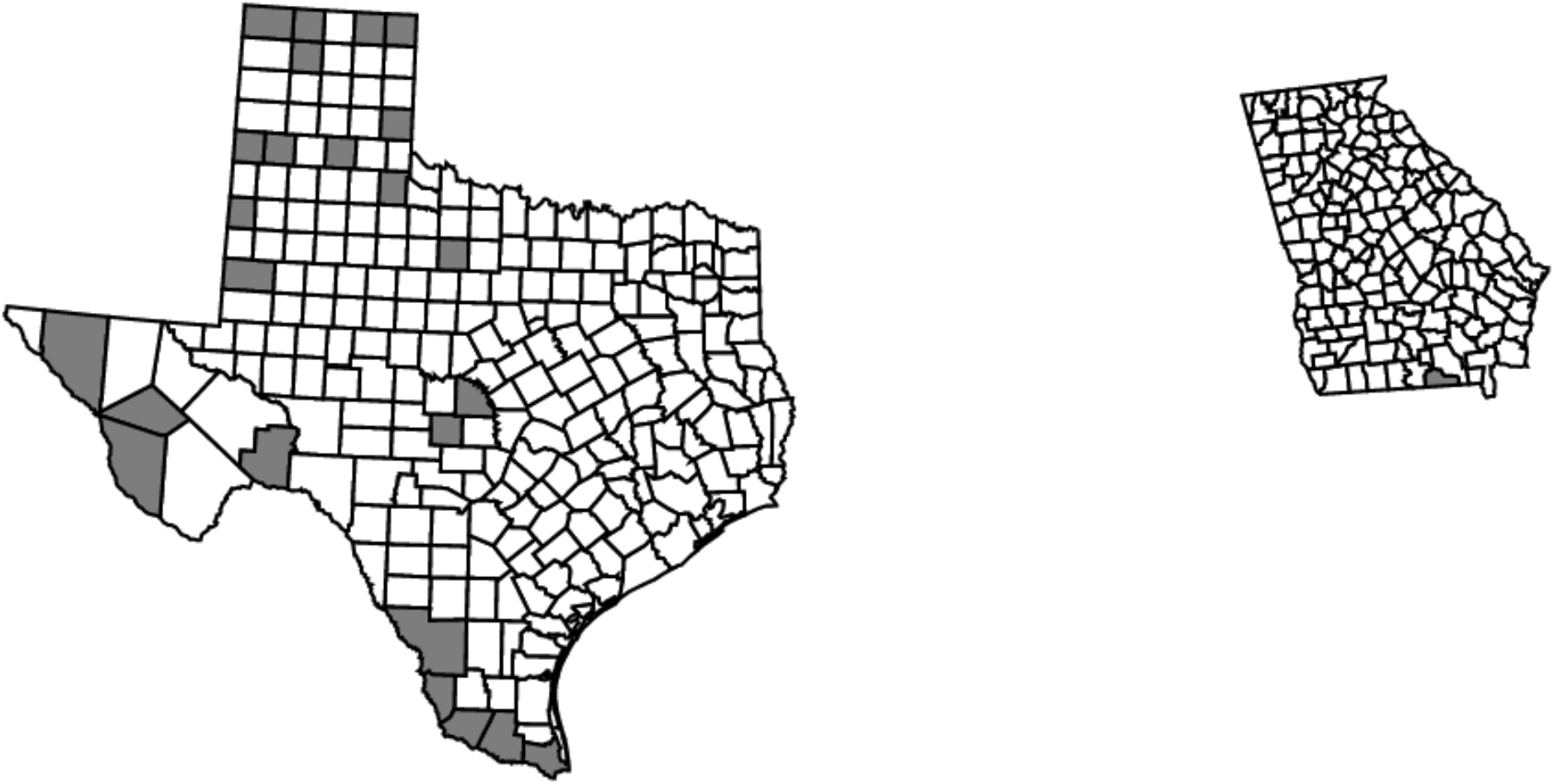
Map of the counties in Texas and Georgia from *Supplementary Table S1*.

**Supplementary Figure S4:**
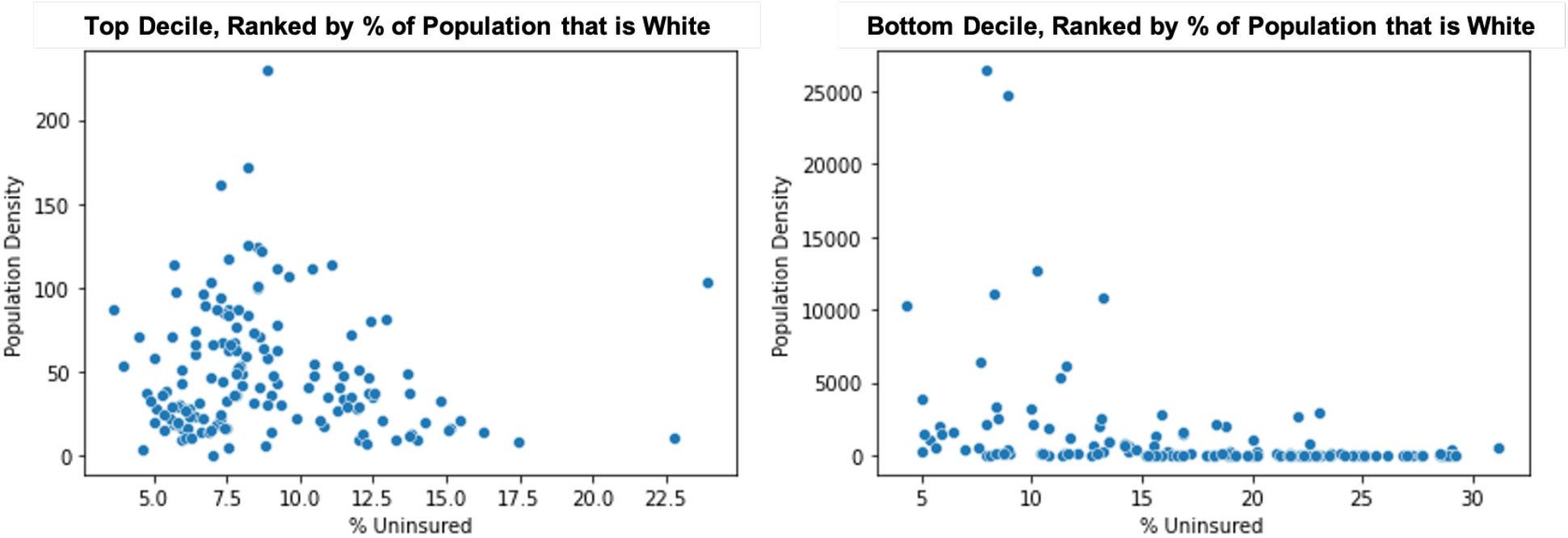
Population density as it relates to percent of the population that is uninsured, in the top decile of counties ranked by percent of population that is white (left) vs. bottom decile (right).

**Supplementary Table S1.** Top 25 US Counties ranked by percent of uninsured population. Information on state, county, percent uninsured, percent vaccinated, increase in COVID-incidence as of March 1, 2021 relative to December 1, 2020, percent rural, population and percent male are provided for each county.

| Uninsured Rank | State | County | % Uninsured | % Vaccinated | COVID incidence increase from Dec 1 to Mar 1 | % Rural | Population | % Male |
| --- | --- | --- | --- | --- | --- | --- | --- | --- |
| 1 | Texas | Gaines County | 33.7 | 3.0 | -20.0 | 63.0 | 21,492 | 50.6 |
| 2 | Texas | Collingsworth County | 32.7 | 8.8 | -10.3 | 100.0 | 2,920 | 49.0 |
| 3 | Texas | Briscoe County | 31.4 | 11.3 | 6.5 | 100.0 | 1,546 | 49.7 |
| 4 | Texas | Hidalgo County | 31.2 | 7.3 | 20.8 | 5.1 | 868,707 | 49.0 |
| 5 | Texas | Dallam County | 30.5 | 16.7 | -1.4 | 23.5 | 7,287 | 52.6 |
| 6 | Texas | Sherman County | 29.6 | 8.1 | 3.3 | 100.0 | 3,022 | 52.2 |
| 7 | Texas | Lipscomb County | 29.5 | 6.0 | -27.8 | 100.0 | 3,233 | 51.1 |
| 8 | Texas | Jeff Davis County | 29.4 | 8.5 | -88.0 | 100.0 | 2,274 | 51.2 |
| 9 | Texas | Zapata County | 29.2 | 4.4 | -47.3 | 23.5 | 14,179 | 49.7 |
| 10 | Texas | Presidio County | 29.0 | 11.6 | -268.5 | 40.5 | 6,704 | 51.3 |
| 11 | Texas | Cameron County | 29.0 | 9.1 | -7.6 | 8.4 | 423,163 | 48.8 |
| 12 | Texas | San Saba County | 28.9 | 1.7 | -16.5 | 49.4 | 6,055 | 54.2 |
| 13 | Texas | Starr County | 28.9 | 5.0 | 7.7 | 23.7 | 64,633 | 48.8 |
| 14 | Texas | Castro County | 28.9 | 8.2 | -37.2 | 45.8 | 7,530 | 51.3 |
| 15 | Texas | Parmer County | 28.7 | 9.3 | -27.1 | 60.0 | 9,605 | 51.7 |
| 16 | Texas | Ochiltree County | 28.6 | 5.4 | -58.0 | 13.9 | 9,836 | 50.2 |
| 17 | Texas | Webb County | 28.6 | 6.2 | -31.1 | 2.6 | 276,652 | 49.2 |
| 18 | Texas | Hudspeth County | 28.5 | 6.3 | -198.5 | 100.0 | 4,886 | 53.1 |
| 19 | Texas | Cochran County | 28.5 | 6.6 | 3.5 | 100.0 | 2,853 | 50.7 |
| 20 | Texas | Mason County | 28.1 | 5.4 | 21.1 | 100.0 | 4,274 | 51.1 |
| 21 | Texas | Terrell County | 27.7 | 21.4 | -154.6 | 100.0 | 776 | 50.4 |
| 22 | Texas | Moore County | 27.7 | 7.3 | -15.8 | 16.8 | 20,940 | 51.9 |
| 23 | Georgia | Echols County | 27.6 | 7.0 | -12.5 | 100.0 | 4,006 | 51.8 |
| 24 | Texas | Cottle County | 27.4 | 4.4 | -28.6 | 100.0 | 1,398 | 48.7 |
| 25 | Texas | Throckmorton County | 27.3 | 18.5 | 0.0 | 100.0 | 1,501 | 47.8 |

